# Development and External Validation of a Machine Learning Model to Predict Restriction from Spirometry

**DOI:** 10.1101/2025.01.02.25319890

**Authors:** Alexander T. Moffett, Aparna Balasubramanian, Meredith C. McCormack, Jaya Aysola, Lyle H. Ungar, Scott D. Halpern, Gary E. Weissman

**Affiliations:** Division of Pulmonary, Allergy, and Critical Care Medicine, Department of Medicine, University of Pennsylvania, Philadelphia, PA, USA; Palliative and Advanced Illness Research (PAIR) Center, University of Pennsylvania, Philadelphia, PA, USA; Leonard Davis Institute of Health Economics, University of Pennsylvania, Philadelphia, PA, USA; Division of Pulmonary and Critical Care Medicine, Johns Hopkins University, Baltimore, MD, USA; Penn Medicine Center for Health Equity Advancement, Office of the Chief Medical Officer, University of Pennsylvania Health System, Philadelphia, PA, USA; Division of General Internal Medicine, Department of Medicine, University of Pennsylvania, Philadelphia, PA, USA; Department of Computer and Information Science, University of Pennsylvania, Philadelphia, PA, USA; Department of Biostatistics, Epidemiology, and Informatics, University of Pennsylvania, Philadelphia, PA, USA; Department of Medical Ethics and Health Policy, University of Pennsylvania, Philadelphia, PA, USA

**Author notes:** Corresponding Author: Alexander T. Moffett Hospital of the University of Pennsylvania Division of Pulmonary, Allergy, and Critical Care Medicine 3400 Spruce Street Philadelphia, PA 19104.

**Keywords:** health equity, machine learning, pulmonary function tests, restriction

## Abstract

**Background:** Though European Respiratory Society and American Thoracic Society (ERS/ATS) guidelines for pulmonary function test (PFT) interpretation recommend the use of the forced vital capacity (FVC) lower limit of normal (LLN) to exclude restriction, recent data suggest that the negative predictive value (NPV) of the FVC LLN is lower than has been accepted, particularly among non-Hispanic Black patients. We sought to develop and externally validate a machine learning (ML) model to predict restriction from spirometry and determine whether its use may improve the accuracy and equity of PFT interpretation.

**Methods:** We included PFTs with both static and dynamic lung volume measurements for patients between 18 and 80 years of age who were tested at pulmonary diagnostic labs within two health systems. We used PFTs from one health system to train logistic regression, random forest, and boosted tree models to predict restriction using demographic, anthropometric, and spirometric data. We used PFTs from the second health system to externally validate these models. The primary measure of model performance was the NPV. Racial equity was assessed by comparing the NPV among non-Hispanic Black and non-Hispanic White patients.

**Findings:** A total of 42 462 PFTs were used for model development and 24 524 for external validation. The prevalence of restriction was 29.8% in the development dataset and 39.6% in the validation dataset. All three ML models outperformed the FVC LLN by a wide margin, both overall and among all demographic subgroups. The overall NPV of the random forest model (88.3%, 95% confidence interval [CI] 87.8% to 88.9%) was significantly greater than that of the FVC LLN (72.7%, 95% CI 72.1% to 73.3%). The NPV of the random forest model was greater than that of the FVC LLN among both non-Hispanic Black (74.6% [95% CI 72.5% to 76.6%] versus 49.5% [95% CI 47.8% to 51.2%]) and non-Hispanic White (90.9% [95% CI 90.3% to 91.5%] versus 79.6% [95% CI 78.9% to 80.3%]) patients.

**Interpretation:** ML models to exclude restriction from spirometry improve the accuracy and equity of PFT interpretation but do not fully eliminate racial differences.

## Introduction

A central purpose of pulmonary function test (PFT) interpretation is to determine the presence or absence of restriction.^1^ European Respiratory Society and American Thoracic Society (ERS/ATS) guidelines for PFT interpretation define restriction by the presence of a total lung capacity (TLC) less than the lower limit of normal (LLN).^2^ According to these guidelines, while the presence of restriction cannot be established from spirometry alone, the absence of restriction can be established on the basis of a normal forced vital capacity (FVC).

To support this recommendation, ERS/ATS guidelines cite a single study from 1999 in which the negative predictive value (NPV) of a normal FVC was estimated at 97.6%.^3^ This study, however, included fewer than two thousand PFTs, was performed at a single pulmonary diagnostic lab, was limited to White patients, and relied on race-specific reference equations that are no longer recommended by ERS/ATS.^4^ We recently reassessed this issue in a diverse, multicenter cohort using race-neutral reference equations and found the NPV of the FVC LLN was 80.5% overall, including 65.2% among non-Hispanic Black patients and 85.9% among non-Hispanic White patients.^5^ This finding suggests that the use of the FVC LLN to exclude restriction may lead to the under-recognition of restriction, particularly among non-Hispanic Black patients.

The potential of machine learning (ML) to improve PFT interpretation has been widely noted,^6–8^ and ML models have been developed to predict the presence of restriction from spirometry.^9–11^ However, the potential of ML to exclude restriction restriction—and to do so equitably across racial groups—remains untested. We therefore developed and externally validated ML models to predict the presence and absence of restriction using spirometric, demographic, and anthropometric data. We then compared the accuracy and equity of the PFT interpretations produced using these ML models with those produced using the FVC LLN to exclude restriction.

## Methods

### Pulmonary Function Tests

For model development, we used PFTs with both static and dynamic lung volume measurements that were performed between 2000 and 2023 at one of three pulmonary diagnostic labs in an academic health system For external validation, we used PFTs with both static and dynamic lung volume measurements that were performed in one of four pulmonary diagnostic labs in a second academic health system. PFTs in both the development and validation datasets were performed in accordance with ERS/ATS guidelines.^12–14^ Static lung volumes were measured with plethysmography in the development dataset and with either plethysmography or helium washout in the validation dataset. For patients with multiple PFTs we included the first PFT performed for each patient. PFTs with missing demographic, anthropometric, or spirometric data were excluded.

PFTs were interpreted in accordance with current ERS/ATS guidelines.^2^ FVC z-scores were calculated using race-neutral GLI Global equations,^15^ while TLC z-scores were calculated using GLI 2019 equations for TLC.^16^ We excluded patients younger than 18 years of age and those older than 80 years of age. A parameter value was interpreted as normal if its z-score was greater than *−*1.645. Restriction was present if the TLC was abnormal.

### Model Development

PFTs in the development dataset were used to train logistic regression, random forest,^17^ and boosted tree^18^ models to predict restriction from demographic, anthropometric, and spirometric data. Age, standing height, weight, sex, forced expiratory volume in 0.5 seconds (FEV_0.5_), FEV_1_, FEV_3_, FEV_6_, FVC, FEV_1_/FVC, forced expiratory flow between 25% and 75% of vital capacity (FEF_25_*_−_*_75_), maximum forced expiratory flow (FEF_Max_), and expiratory time were selected as predictors on the basis of prior clinical knowledge. Raw values were used for all predictors. Training was performed using 10-fold cross validation, repeated 5 times. Hyperparameter tuning was performed using complete grid search, to optimize the area under the receiver operating characteristic curve (AUC-ROC).

### Model Validation

The NPV was the primary measure of model performance, with sensitivity, specificity, and positive predictive value also assessed. As our ML models predict the probability of restriction, thresholds were needed to transform these continuous probabilities into binary predictions, so as to calculate these threshold-dependent measures of model performance. A threshold was selected for each model so as to maximize the sum of the sensitivity and specificity in the development dataset. In addition to the NPV, threshold-dependent model performance was further assessed by the percentage of PFTs in which the need for static lung volume measurement was excluded due to the prediction of a normal TLC.

Model performance was assessed in a threshold-independent manner using the AUC-ROC, the area under the precision-recall curve (AUC-PR), the integrated calibration index (ICI), and the scaled Brier score (SBS). Both the AUC-ROC and the AUC-PR provide measures of model discrimination, while the ICI provides a measure of model calibration.^19^ The SBS provides a combined measure of both discrimination and calibration.^20^ We calculated 95% confidence intervals for these estimates using 1000 bootstrapped samples.

Internal validation was performed by applying the ML models trained with the development dataset to the development dataset, with the mean cross-validated performance reported to mitigate for the optimism associated with applying a model to the data used to train it. External validation was performed by applying the ML models trained with the development dataset to the validation dataset.^21^

To assess model equity, model performance was compared between non-Hispanic White and non-Hispanic Black patients. Racial and ethnic data were self-reported. The combined use of race and ethnicity reflects the historical use of reference equations for spirometry in which Hispanic patients were interpreted as having different baseline pulmonary function than White and Black patients.^22^

We used Shapley values to explain the predictions of the ML models.^23^ For each model, we calculated Shapley values for 1 000 samples of 100 observations and calculated the mean absolute value of the Shapley value for each predictor across these samples, comparing these values to determine the relative importance of the different predictors to each model.

A decision curve analysis was performed to compare the clinical value of the ML models with that of the FVC LLN.^24^

### Statistical Computing

The analysis was performed using the R programming language for statistical computing.^25^ The ranger^26^ package used for the random forest model and XGBoost^27^ was used for the boosted tree model. Shapley values were calculated using shapr.^28^ All code is open source (https://github.com/weissman-lab/restriction) and all model objects are freely available.

The University of Pennsylvania and Johns Hopkins Hospital Institutional Review Boards approved this study. The study was performed in accordance with the transparent reporting of a multivariable prediction model for individual prognosis or diagnosis (TRIPOD+AI) reporting standards (**Table S1**).^29^

## Results

The development and validation datasets included 42 462 and 24 524 PFTs, respectively (**Table 1**, **Figure S1**). Most patients in both the development and validation datasets were women and were non-Hispanic White or non-Hispanic Black. Patients in the development dataset had lower median FEV_1_ and FVC z-scores (*−*1.3 and *−*1.0, respectively) than patients in the validation dataset (*−*0.8 and *−*0.5), while patients in the validation dataset had lower median TLC z-scores (*−*1.3) than those in the development dataset (*−*0.8). The prevalence of restriction was higher in the validation dataset (39.6%) than in the development dataset (29.8%). Patient characteristics varied by race and ethnicity in both the development (**Table S2**) and validation (**Table S3**) datasets, with non-Hispanic Black patients having lower median FEV_1_, FVC, and TLC z-scores than non-Hispanic White patients, along with a higher prevalence of restriction.

**Table 1:** Patient Characteristics.

|  | Health System 1:<br>Development<br>Dataset<br>(*n* = 42 462) |  | Health System 2:<br>Validation<br>Dataset<br>(*n* = 24 524) |  |
| --- | --- | --- | --- | --- |
| Demographics |  |  |  |  |
| Age |  |  |  |  |
| 18–40 | 7 160 | (16.9) | 3 348 | (13.7) |
| 41–64 | 21 889 | (51.5) | 11 852 | (48.3) |
| 65–80 | 13 413 | (31.6) | 9 324 | (38.0) |
| Sex |  |  |  |  |
| Male | 17 598 | (41.4) | 10 504 | (42.8) |
| Female | 24 864 | (58.6) | 14 020 | (57.2) |
| Race and Ethnicity |  |  |  |  |
| Asian | 505 | (1.2) | 95 | (0.4) |
| Hispanic | 524 | (1.2) | 21 | (0.1) |
| Non-Hispanic Black | 13 867 | (32.7) | 5 584 | (22.8) |
| Non-Hispanic White | 26 261 | (61.8) | 14 280 | (58.2) |
| Other | 1 305 | (3.1) | 4 544 | (18.5) |
| Spirometry |  |  |  |  |
| FEV<sub>1</sub>, z-score | −1.3 | (1.9) | −0.8 | (1.9) |
| FVC, z-score | −1.0 | (1.8) | −0.5 | (1.8) |
| FEV<sub>1</sub>/FVC, z-score | −0.5 | (1.8) | −0.5 | (1.5) |
| Lung Volumes |  |  |  |  |
| TLC, z-score | −0.8 | (2.0) | −1.3 | (1.9) |
| ERS/ATS Classification |  |  |  |  |
| Normal | 18 626 | (43.9) | 11 035 | (45.0) |
| Non-Specific | 2 800 | (6.6) | 424 | (1.7) |
| Obstructive | 8 383 | (19.7) | 3 351 | (13.7) |
| Restrictive | 11 454 | (27.0) | 8 266 | (33.7) |
| Mixed | 1 199 | (2.8) | 1 448 | (5.9) |
| ERS/ATS Severity |  |  |  |  |
| Normal | 25 286 | (59.5) | 17 315 | (70.6) |
| Mild | 8 239 | (19.4) | 3 845 | (15.7) |
| Moderate | 7 357 | (17.3) | 2 963 | (12.1) |
| Severe | 1 580 | (3.7) | 401 | (1.6) |

Definition of abbreviations: ATS = American Thoracic Society; ERS = European Respiratory Society; FEV<sub>1</sub> = forced expiratory volume in 1 second; FVC = forced vital capacity; GLI = Global Lung Function Initiative; IQR = interquartile range; TLC = total lung capacity.

The highest overall performance was seen with the random forest model. As applied to the validation dataset, this model had an NPV of 88.3% (95% CI 87.8% to 88.9%), while the FVC LLN had an NPV of 72.7% (95% CI 72.1% to 73.3%) (**Table 2**). The random forest model excluded the need for static lung volume measurements in 56.7% (95% CI 56.1% to 57.3%) of tests, while the FVC LLN excluded the need for these measurements in 78.4% (95% CI 77.9% to 78.9%) of tests. The sensitivity of the random forest model was 83.3% (95% CI 82.5% to 84.0%), while the sensitivity of the FVC LLN was 46.0% (95% CI 45.0% to 47.0%). The specificity of the random forest model was 82.9% (95% CI 82.4% to 83.5%), while the specificity of the FVC LLN was 94.4% (95% CI 94.0% to 94.7%).

**Table 2:** Model Performance as Applied to the Validation Dataset.

| Model | Threshold-Dependent Model Performance |  |  |  | Threshold-Independent Model Performance |  |  |  |
| --- | --- | --- | --- | --- | --- | --- | --- | --- |
|  | Sensitivity (%) | Specificity (%) | NPV (%) | PPV (%) | AUC-ROC | AUC-PR | ICI | SBS |
| FVC < LLN | 46.0 (45.0–47.0) | 94.4 (94.0–94.7) | 72.7 (72.1–73.3) | 84.3 (83.3–85.3) | — | — | — | — |
| Random Forest | 83.3 (82.5–84.0) | 82.9 (82.4–83.5) | 88.3 (87.8–88.9) | 76.2 (75.4–77.0) | 0.91 (0.91–0.92) | 0.88 (0.87–0.88) | 0.17 (0.17–0.18) | 0.32 (0.30–0.33) |
| Boosted Tree | 83.0 (82.3–83.8) | 83.3 (82.7–83.9) | 88.2 (87.7–88.8) | 76.6 (75.7–77.3) | 0.92 (0.91–0.92) | 0.88 (0.87–0.89) | 0.17 (0.17–0.18) | 0.32 (0.31–0.33) |
| Logistic Regression | 82.4 (81.6–83.1) | 83.5 (83.0–84.1) | 87.8 (87.3–88.4) | 76.6 (75.8–77.0) | 0.91 (0.91–0.91) | 0.87 (0.87–0.88) | 0.19 (0.19–0.20) | 0.25 (0.24–0.27) |

Definition of abbreviations: AUC-PR = area under the precision-recall curve; AUC-ROC = area under the receiver operating characteristic curve; FVC = forced vital capacity; ICI = integrated calibration index; LLN = lower limit of normal; NPV = negative predictive value; PPV = positive predictive value; SBS = scaled Brier score.

The random forest model possessed excellent discrimination but was poorly calibrated (**Table 2**). The AUC-ROC of random forest model was 0.91 (95% CI 0.91 to 0.92), while the AUC-PR was 0.88 (95% CI 0.87 to 0.88) (**Figures 1a**). The ICI of the random forest model was 0.17 (95% CI 0.17 to 0.18), with the observed frequency of restriction consistently higher than the predicted probability of restriction (**Figure 1b**). The SBS of the random forest model was 0.32 (95% CI 0.30 to 0.32). There was a significant decline in performance in moving from the development to the validation dataset (**Table S4**).

**Figure 1.**
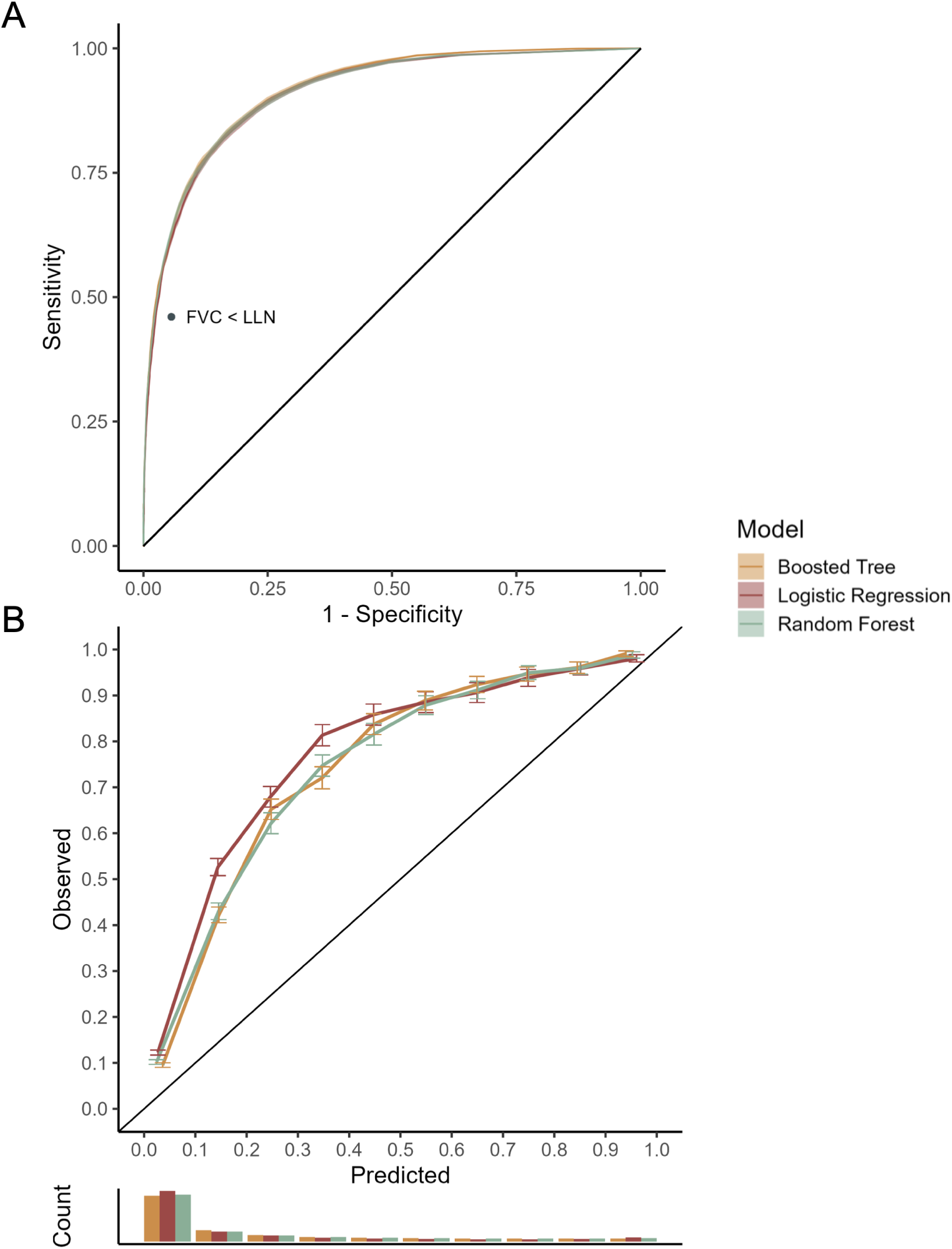
Model discrimination and calibration. Receiver operating characteristic curves (A) represent the discrimination of logistic regression, random forest, and boosted tree models for the prediction of restriction from spirometry, as applied to the validation data set. Calibration plots (B) represent the calibration of logistic regression, random forest, and boosted tree models for the prediction of restriction from spirometry, as applied to the validation data set.

Model performance differed between non-Hispanic Black and non-Hispanic White patients (**Table 3, S5**). The NPV of the FVC LLN was 49.5% (95% CI 47.8% to 51.2%) among non-Hispanic Black patients and 79.6% (95% CI 78.9% to 80.3%) among non-Hispanic White patients. The NPV of the random forest model was 74.6% (95% CI 72.5% to 76.6%) among non-Hispanic Black patients and 90.9% (95% CI 90.3% to 91.5%) among non-Hispanic White patients. The ratio of the NPV among non-Hispanic Blacks and non-Hispanic Whites was greater with the random forest model (82.1%, 95% CI 79.6% to 84.4%) than with the FVC LLN (62.2%, 95% CI 60.1% to 64.3%). Performance was similar between male and female patients and between patients younger and older than 65 years of age (**Table S6, S7**).

**Table 3:** Negative Predictive Value by Race in the Validation Dataset.

| Model | NPV (%) |  | Ratio of Lowest<br>to Highest NPV (%) |
| --- | --- | --- | --- |
|  | Non-Hispanic<br>Black Patients | Non-Hispanic<br>White Patients |  |
| FVC < LLN | 49.5 (47.8 to 51.2) | 79.6 (78.9 to 80.3) | 62.2 (60.1 to 64.3) |
| Random Forest | 74.6 (72.5 to 76.6) | 90.9 (90.3 to 91.5) | 82.1 (79.6 to 84.4) |
| Boosted Tree | 74.5 (72.6 to 76.5) | 90.9 (90.2 to 91.4) | 82.0 (79.7 to 84.3) |
| Logistic Regression | 72.6 (70.6 to 74.9) | 90.6 (90.0 to 91.1) | 80.2 (77.9 to 82.6) |

Definition of abbreviations: FVC = forced vital capacity; LLN = lower limit of normal; NPV = negative predictive value.

Much of the improvement in ML model performance was seen in patients with an FVC near the LLN (**Figure S2**). For PFTs with an FVC z-score in the interval [*−*1.645, *−*1.445), the NPV of a normal FVC was 26.5% (95% CI 23.9% to 29.3%), while the NPV of the random forest model in this interval was 72.0% (95% CI 63.3% to 79.7%). The NPV of the FVC LLN was less than that of the boosted tree model for FVC z-scores in intervals *< −*0.045 and was equal to that of the boosted tree model for FVC z-scores in intervals *≥ −*0.045. The random forest model excluded restriction from significantly fewer PFTs with an FVC z-score *<* 0.155 (**Figure S3**).

The ML models differed with respect to the relative importance of the predictors (**Figures S4–S6**). As measured by the mean absolute value of the Shapley values, the FVC was the most important predictor for the random forest model, while height was the most important predictor for the boosted tree model, and FEV_1_/FVC was the most important predictor for the logistic regression model.

In the decision curve analysis, for thresholds less than 0.1, the measurement of static lung volumes in all patients was preferred to the use of either the random forest model or the FVC LLN (**Figure 2**). For thresholds between 0.10 and 0.58 and between 0.77 and 1.0, the use of the random forest model was preferred both to the measurement of static lung volumes in all patients and to the use of the FVC LLN. The FVC LLN was preferred for thresholds between 0.59 and 0.76.

**Figure 2.**
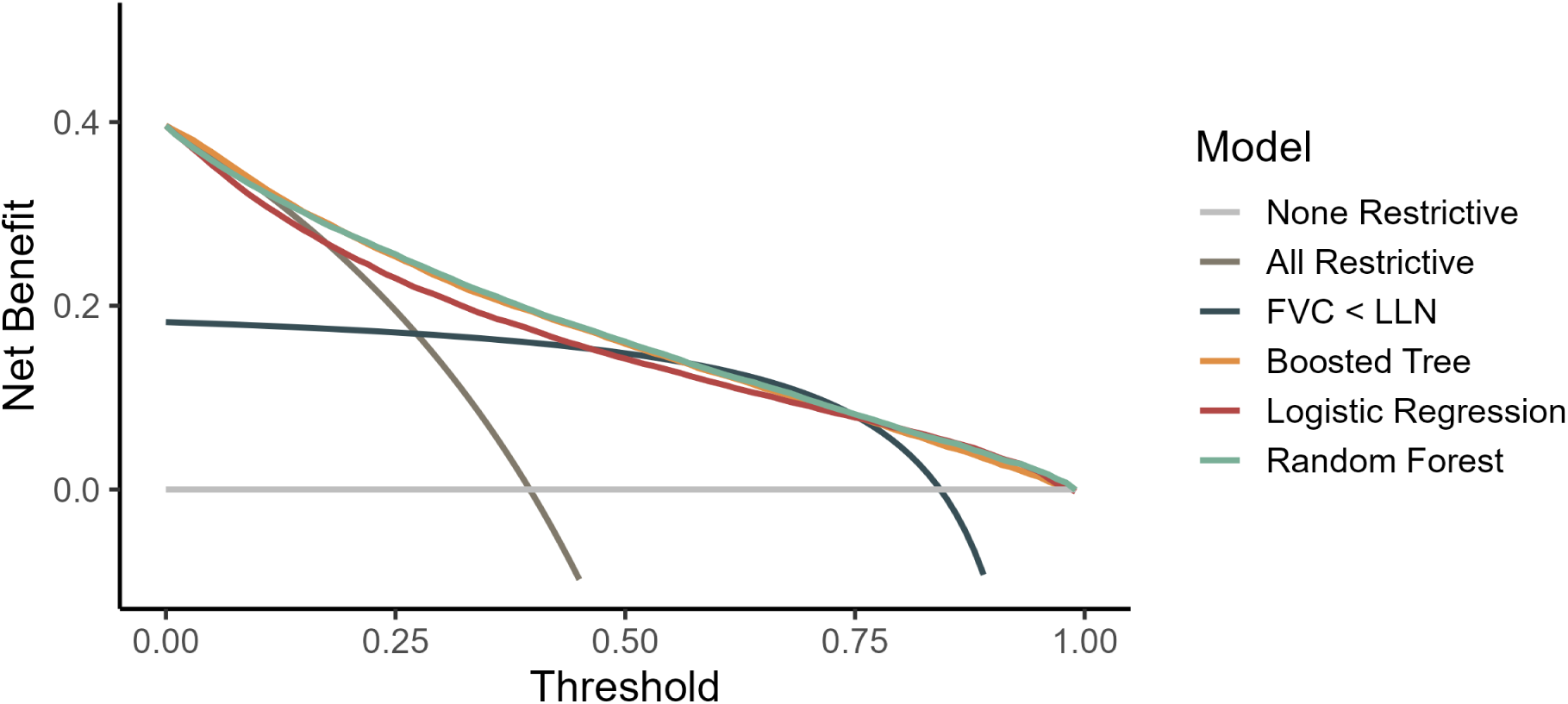
Decision curve analysis. Decision curves show the net benefit associated with different models at different threshold values, as applied to the validation data set. The dashed gray line shows the net benefit per patient under the assumption that no PFTs are restrictive. The gray line shows the net benefit per patient under the assumption that all PFTs are restrictive. The black line shows the net benefit per patient under the assumption that PFTs with an FVC *<* LLN are restrictive.The orange, red, and green curves show the net benefit per patient under the boosted tree, logistic regression, and random forest models, respectively. FVC = forced vital capacity; LLN = lower limit of normal; PFTs = pulmonary function tests.

## Discussion

We developed and externally validated three ML models to predict restriction from spirometric, anthropometric, and demographic data. Performance was similar across the three models, with the greatest overall performance seen in the random forest model. We found that all ML models were more accurate and equitable than the FVC LLN in excluding restriction from spirometry.

ERS/ATS guidelines are predicated on the idea that a normal FVC can be used to predict a normal TLC with near-perfect accuracy.^3^ Recent results, however, suggest that when applied to diverse patient populations with modern race-neutral reference equations, the accuracy of this prediction is significantly lower, particularly among non-Hispanic Black patients.^5^ In this study we found that accuracy can be improved by using a ML model to predict restriction from spirometry. In our validation dataset we found that a random forest model possessed an NPV of 88% compared to an NPV of 73% on the part of the FVC LLN. There are several reasons why the ML model outperformed the FVC LLN. The ML models developed in this study include multiple spirometric parameters—the FEV_0.5_, FEV_1_, FEV_3_, FEV_6_, FEV_1_/FVC, FEF_25–75_, FEF_Max_, and expiratory time—in addition to the FVC. And rather than consider only a binary representation of these predictors—whether the predictor was above or below the LLN—our approach to model development instead allowed for a range of potential representations of these predictors, along with the complex nonlinear relationships between them. The inclusion of additional spirometric data and the consideration of a larger space of potential models led to the development of more accurate clinical prediction models.

While the random forest model was significantly more accurate than the FVC LLN in excluding restriction, model selection should be based upon more than just accuracy. The random forest model excluded restriction from far fewer PFTs than did the FVC LLN and its adoption would significantly increase the number of PFTs for which static lung volumes are recommended. What is missing from the current recommended use of the FVC LLN to exclude restriction is an assessment of the preferences of patients and providers regarding the cost of a false negative result—the misclassification of restriction as normal spirometry— relative to the cost of a false positive result—the measurement of static lung volumes in a patient with normal spirometry.^30^ In the absence of a formal decision analysis, our decision curve analysis offers some insight into the net benefit of different models across a range of different decision thresholds. In this analysis we found that the random forest model outperformed the FVC LLN for almost all thresholds, with the FVC LLN preferred only between thresholds of 0.59 and 0.76. That is, the FVC LLN was preferred only when the measurement of static lung volumes in a patient with a normal TLC was thought worse than missing a case of restriction. We would expect that patients and providers would regard a missed instance of restriction as significantly worse given the minimal cost associated with static lung volume measurement. Indeed, it is notable that for thresholds of less than 0.10, the measurement of static lung volumes in all patients was preferred to the use of either the random forest model or the FVC LLN to predict the need for static lung volume measurement.

In addition to improving the accuracy of PFT interpretation, the boosted tree model also improved the equity of PFT interpretation. Among non-Hispanic Black patients, the NPV of the FVC LLN was less than 50% while the NPV of the boosted tree model was close to 75%. The ratio of the NPV among non-Hispanic Black patients to the NPV among non-Hispanic White patients increased from 62% with the FVC LLN to 82% with the boosted tree model. The FVC LLN performed poorly when applied to patients with lower FVC z-scores and the FVC z-scores of non-Hispanic Black patients were significantly lower than those of non-Hispanic White patients, leading to substantial inequity in model performance by race. An improvement in equity is seen with the adoption of a more sophisticated model, in which performance is less dependent upon the FVC z-score. While the adoption of ML is often framed in terms of a tradeoff between accuracy and equity, in this case the ML model yielded a significant improvement along both dimensions. This finding highlights the way in which guidelines employed in current clinical practice, though developed without ML, can still contribute to inequity. The equity concerns that have been rightly posed in response to the development of ML models should be applied to current guidelines as well.

This study has several strengths. First, we developed multiple ML models and assessed model performance with multiple performance measures, including a decision curve analysis. While these models were found to function quite differently, their similar performance supports the robustness of our results. Second, we externally validated our models, developing them with PFT data from one academic health system and then assessing the performance of these models when applied to PFT data from a second academic health system, and found that the performance of the ML models exceeded that of the FVC LLN in both the development and validation datasets. Third, in addition to assessing the overall performance of our models, we further assessed the potential impact of these models on the equity of PFT interpretation by comparing their performance among non-Hispanic Black and non-Hispanic White patients, as well as among male and female patients and older and younger patients. Fourth, the code used in model development is open source and the models themselves are publicly accessible, allowing other researchers to build similar models using local data and compare the performance of these models to that reported in this study.

This study also has important limitations. First, as the ML models were developed and validated with PFT data from academic health systems—in which the prevalence of restriction is likely higher than in pulmonary diagnostic labs in the community—the potential for these models to overestimate the probability of restriction should be considered. Indeed, it is notable that though the ML models in this study were developed with PFT data from an academic health system, the models were nonetheless poorly calibrated when externally validated with data from a second academic health system. Second, as our study was limited to PFTs in which both static and dynamic lung volumes were measured, there is the potential for ascertainment bias. This could affect the absolute values of the performance characteristics of all models, but because all were run on the same patients, should not affect the major conclusion that the ML models were superior. Third, while our ML models included additional spirometric data not considered by ERS/ATS guidelines, additional spirometric data could be included in the form of the complete flow-volume loop. Future models, applying deep learning to these data, may see further improvements in performance. Fourth, due to the limited racial and ethnic diversity within our validation dataset, our equity assessment only compared model performance in non-Hispanic Black and non-Hispanic White patients. Future efforts to continue to externally validate these models should consider their equity implications for populations from other racial and ethnic groups as well as populations of multi-racial and ethnic origins. Fifth, our equity assessment was limited to the comparison of model performance and did not include feedback from patients and providers. It is notable that though the ML models were more accurate than the FVC LLN in excluding restriction among non-Hispanic Black patients, significant inequity nonetheless remained. Rather than use ML to provide a technical solution to the inequity associated with current guidelines, more fundamental changes in our approach to PFT interpretation may be needed.

## Conclusion

We developed and externally validated ML models to predict restriction from spirometry. We found that using a ML model—rather than the FVC LLN—to exclude restriction resulted in an improvement in both the accuracy and the equity of PFT interpretation.

## Data Availability

All data produced in the present study are available upon reasonable request to the authors.

https://github.com/weissmanlab/restriction

## Contributions

ATM participated in study design and analysis and drafted and revised the manuscript. AB, MCM, JA, LHU, SDH and GEW participated in study design and analysis and revised the manuscript. All authors have read and approved the manuscript.

