## Supplement for "Development and External Validation of a Machine Learning Model to Predict Restriction from Spirometry"

Tables

**Table S1**. TRIPOD+A1 Checklist

**Table S2**. Patient Characteristics in the Development Dataset by Race and Ethnicity

**Table S3**. Patient Characteristics in the Validation Dataset by Race and Ethnicity

**Table S4**. Model Performance in the Development and Validation Datasets

**Table S5**. Model Performance in the Validation Dataset by Race and Ethnicity

**Table S6**. Model Performance in the Validation Dataset by Age Group

**Table S7**. Model Performance in the Validation Dataset by Sex

Figures

**Figure S1**. Flow Diagram for Inclusion in the Development Dataset

**Figure S2**. Forced Vital Capacity Z-Score and Negative Predictive Value

**Figure S3**. Forced Vital Capacity Z-Score and the Exclusion of Restriction

**Figure S4**. Boosted Tree Model Shapley Values

**Figure S5**. Logistic Regression Model Shapley Values

**Figure S6**. Random Forest Model Shapley Values

**Table S1**. TRIPOD+AI Checklist

| Section/Item | Item | Checklist Item | Location |
|---|---|---|---|
| | | *Title* | |
| Title | 1 | Identify the study as developing or evaluating the performance of a multivariable prediction model, the target population, and the outcome to be predicted | Page 1 |
| | | *Abstract* | |
| Background | 2a | Provide a brief explanation of the healthcare context and rationale for developing or evaluating the performance of all models | Page 1 |
| Objective | 2b | Specify the study objectives, including whether the study describes model development, evaluation, or both | Page 1 |
| | 2c | Describe the sources of data | Page 1 |
| | 2d | Describe the eligibility criteria and setting where the data were collected | Page 1 |
| Methods | 2e | Specify the outcome to be predicted by the model, including time horizon of predictions in case of prognostic models | Page 1 |
| | 2f | Specify the type of model, a summary of the model-building steps, and the method for internal validation | Page 1 |
| | 2g | Specify the measures used to assess model performance (eg, discrimination, calibration, clinical utility) | Page 1 |
| | 2h | Report the number of participants and outcome events | Page 2 |
| Results | 2i | Summarise the predictors in the final model | Page 1 |
| | 2j | Report model performance estimates (with confidence intervals) | Page 2 |
| Discussion | 2k | Give an overall interpretation of the main results | Page 2 |
| Registration | 2l | Give the registration number and name of the registry or repository | N/A |
| | | *Introduction* | |
| | 3a | Explain the healthcare context (including whether diagnostic or prognostic) and rationale for developing or evaluating the prediction model, including references to existing models | Page 3 |
| Background | 3b | Describe the target population and the intended purpose of the prediction model in the context of the care pathway, including its intended users (e.g., healthcare professionals, patients, public) | Page 3 |
| | 3c | Describe any known health inequalities between sociodemographic groups. | Page 3 |
| Objectives | 4 | Specify the study objectives, including whether the study describes the development or validation of a prediction model (or both) | Page 4 |
| | | *Methods* | |
| Data | 5a | Describe the sources of data separately for the development and evaluation datasets (e.g., randomised trial, cohort, routine care or registry data), the rationale for using these data, and representativeness of the data | Page 5 |
| | 5b | Specify the dates of the collected participant data, including start and end of participant accrual; and, if applicable, end of follow-up | Page 5 |
| | 6a | Specify key elements of the study setting (e.g., primary care, secondary care, general population) including the number and location of centres | Page 5 |
| Participants | 6b | Describe the eligibility criteria for study participants | Page 5 |
| | 6c | Give details of any treatments received, and how they were handled during model development or evaluation, if relevant | N/A |
| Data Preparation | 7 | Describe any data pre-processing and quality checking, including whether this was similar across relevant sociodemographic groups | Figure S1 |
| Outcome | 8a | Clearly define the outcome that is being predicted and the time horizon, including how and when assessed, the rationale for choosing this outcome, and whether the method of outcome assessment is consistent across sociodemographic groups | Page 5 |
| | 8b | If outcome assessment requires subjective interpretation, describe the qualifications and demographic characteristics of the outcome assessors | N/A |
| | 8c | Report any actions to blind assessment of the outcome to be predicted | N/A |

**Table S1 (Continued)**. TRIPOD+AI Checklist

| Section/Item | Item | Recommendation | Location |
|---|---|---|---|
| | | *Methods* | |
| Predictors | 9a | Describe the choice of initial predictors (e.g., literature, previous models, all available predictors) and any pre-selection of predictors before model building | Page 5 |
| | 9b | Clearly define all predictors, including how and when they were measured (and any actions to blind assessment of predictors for the outcome and other predictors) | Page 5 |
| | 9c | If predictor measurement requires subjective interpretation, describe the qualifications and demographic characteristics of the predictor assessors | N/A |
| Sample Size | 10 | Explain how the study size was arrived at (separately for development and evaluation), and justify that the study size was sufficient to answer the research question. Include details of any sample size calculation | Page 5 |
| Missing Data | 11 | Describe how missing data were handled. Provide reasons for omitting any data | Figure S1 |
| Analytical Methods | 12a | Describe how the data were used (e.g., for development and evaluation of model performance) in the analysis, including whether the data were partitioned, considering any sample size requirements | Page 6 |
| | 12b | Depending on the type of model, describe how predictors were handled in the analyses (functional form, rescaling, transformation, or any standardisation) | Page 6 |
| | 12c | Specify the type of model, rationale, all model-building steps, including any hyperparameter tuning, and method for internal validation | Page Page 6 |
| | 12d | Describe if and how any heterogeneity in estimates of model parameter values and model performance was handled and quantified across clusters (e.g., hospitals, countries) | N/A |
| | 12e | Specify all measures and plots used (and their rationale) to evaluate model performance (e.g., discrimination, calibration, clinical utility) and, if relevant, to compare multiple models | Page 6 |
| | 12f | Describe any model updating (e.g., recalibration) arising from the model evaluation, either overall or for particular sociodemographic groups or settings | N/A |
| | 12g | For model evaluation, describe how the model predictions were calculated (e.g., formula, code, object, application programming interface) | Page 7 |
| Class Imbalance | 13 | If class imbalance methods were used, state why and how this was done, and any subsequent methods to recalibrate the model or the model predictions | N/A |
| Model Output | 15 | Specify the output of the prediction model (e.g., probabilities, classification). Provide details and rationale for any classification and how the thresholds were identified | Page 6 |
| Training versus Evaluation | 16 | Identify any differences between the development and evaluation data in healthcare setting, eligibility criteria, outcome, and predictors | Table 1 |
| Ethical Approval | 17 | Name the institutional research board or ethics committee that approved the study and describe the participant-informed consent or the ethics committee waiver of informed consent | Page 7 |
| | | *Open Science* | |
| Funding | 18a | Give the source of funding and the role of the funders for the present study | Title Page |
| | 18b | Declare any conflicts of interest and financial disclosures for all authors | Title Page |
| | 18c | Indicate where the study protocol can be accessed or state that a protocol was not prepared | N/A |
| | 18d | Provide registration information for the study, including register name and registration number, or state that the study was not registered | N/A |
| | 18e | Provide details of the availability of the study data | N/A |
| | 18f | Provide details of the availability of the analytical code | Page 7 |
| | | *Patient & Public Involvement* | |
| Patient & Public Involvement | 19 | Provide details of any patient and public involvement during the design, conduct, reporting, interpretation, or dissemination of the study or state no involvement | N/A |

**Table S1 (Continued)**. TRIPOD+AI Checklist

| **Section/Item** | **Item** | **Recommendation** | **Location** |
|---|---|---|---|
| | | *Results* | |
| Participants | 20a | Describe the flow of participants through the study, including the number of participants with and without the outcome and, if applicable, a summary of the follow-up time | Table 1 |
| | 20b | Report the characteristics overall and, where applicable, for each data source or setting, including the key dates, key predictors (including demographics), treatments received, sample size, number of outcome events, follow-up time, and amount of missing data. Report any differences across key demographic groups. | Tables 1, S2, S3 |
| | 20c | For model evaluation, show a comparison with the development data of the distribution of important predictors (demographics, predictors, and outcome) | Table 1 |
| Model Development | 21 | Specify the number of participants and outcome events in each analysis (e.g., for model development, hyperparameter tuning, model evaluation) | Table 1 |
| Model Specification | 22 | Provide details of the full prediction model (e.g., formula, code, object, application programming interface) to allow predictions in new individuals and to enable third-party evaluation and implementation, including any restrictions to access or re-use (e.g., freely available, proprietary) | Page 7 |
| Model Performance | 23a | Report model performance estimates with confidence intervals, including for any key subgroups | Table 2 |
| | 23b | If examined, report results of any heterogeneity in model performance across clusters. | N/A |
| Model Updating | 24 | Report the results from any model updating, including the updated model and subsequent performance | N/A |
| | | *Discussion* | |
| Interpretation | 25 | Give an overall interpretation of the main results, including issues of fairness in the context of the objectives and previous studies | Page 11 |
| Limitations | 26 | Discuss any limitations of the study (such as a non-representative sample, sample size, overfitting, missing data) and their effects on any biases, statistical uncertainty, and generalizability | Page 13 |
| Usability of the Model | 27a | Describe how poor quality or unavailable input data (e.g., predictor values) should be assessed and handled when implementing the prediction model | N/A |
| | 27b | Specify whether users will be required to interact in the handling of the input data or use of the model, and what level of expertise is required of users | Page 13 |
| | 27c | Discuss any next steps for future research, with a specific view to applicability and generalizability of the model | Page 13 |

**Table S2**. Patient Characteristics in the Development Dataset by Race and Ethnicity

| | **Asian** ($n$ = 505) | **Hispanic** ($n$ = 524) | **Non-Hispanic Black** ($n$ = 13 867) | **Non-Hispanic White** ($n$ = 26 261) | **Other** ($n$ = 1 305) |
|---|---|---|---|---|---|
| **Age** | | | | | |
| 18–14 | 137 (27.1) | 154 (29.4) | 2574 (18.6) | 4000 (15.2) | 295 (22.6) |
| 41–64 | 210 (41.6) | 272 (51.9) | 7907 (57.0) | 12849 (48.9) | 651 (49.9) |
| 65–80 | 158 (31.3) | 98 (18.7) | 3386 (24.4) | 9412 (35.8) | 359 (27.5) |
| **Sex** | | | | | |
| Men | 223 (44.2) | 212 (40.5) | 4037 (29.1) | 12538 (47.7) | 588 (45.1) |
| Women | 282 (55.8) | 312 (59.5) | 9830 (70.9) | 13723 (52.3) | 717 (54.9) |
| **Dynamic Lung Volumes, z-score** | | | | | |
| $FEV_1$ | −0.9 (1.6) | −1.1 (1.9) | −1.8 (1.5) | −1.0 (2.0) | −1.2 (1.9) |
| FVC | −0.8 (1.7) | −1.0 (1.6) | −1.6 (1.4) | −0.6 (1.8) | −0.9 (1.8) |
| $FEV_1$/FVC | −0.2 (1.4) | −0.1 (1.4) | −0.3 (1.9) | −0.6 (1.8) | −0.2 (1.6) |
| **Static Lung Volumes, z-score** | | | | | |
| TLC | −0.7 (1.6) | −0.9 (1.9) | −1.5 (1.7) | −0.4 (1.9) | −1.0 (2.1) |
| **Interpretation** | | | | | |
| Normal | 287 (56.8) | 271 (51.7) | 3889 (28.0) | 13584 (51.7) | 595 (45.6) |
| Non-Specific | 28 (5.5) | 38 (7.3) | 1360 (9.8) | 1304 (5.0) | 70 (5.4) |
| Obstructive | 61 (12.1) | 66 (12.6) | 2239 (16.1) | 5825 (22.2) | 192 (14.7) |
| Restrictive | 121 (24.0) | 140 (26.7) | 5740 (41.4) | 5043 (19.2) | 410 (31.4) |
| Mixed | 8 (1.6) | 9 (3.0) | 639 (4.6) | 505 (1.9) | 38 (2.9) |
| **Severity** | | | | | |
| Normal | 369 (73.1) | 348 (66.4) | 6258 (45.1) | 17491 (66.6) | 820 (62.8) |
| Mild | 75 (14.9) | 84 (16.0) | 3785 (27.3) | 4024 (15.3) | 271 (20.8) |
| Moderate | 60 (11.9) | 82 (15.6) | 3320 (23.9) | 3719 (14.2) | 176 (13.5) |
| Severe | 1 (0.2) | 10 (1.9) | 504 (3.6) | 1027 (3.9) | 38 (2.9) |

Values are median (IQR) for continuous variables and count (percentage) for categorical variables. Dynamic lung volume z-scores are calculated using GLI Global reference equations, while dynamic lung volume z-scores are calculated using GLI 2019 reference equations. Abbreviations: COPD = chronic obstructive pulmonary disease; $FEV_1$ = forced expiratory volume in 1 second; FVC = forced vital capacity; GLI = Global Lung Function Initiative; ILD = interstitial lung disease; LLN = lower limit of normal; TLC = total lung capacity.

**Table S3**. Patient Characteristics in the Validation Dataset by Race and Ethnicity

| | **Asian** ($n = 95$) | **Hispanic** ($n = 21$) | **Non-Hispanic Black** ($n = 5584$) | **Non-Hispanic White** ($n = 14280$) | **Other** ($n = 4544$) |
|---|---|---|---|---|---|
| **Age, years** | | | | | |
| 18–40 | 21 (22.1) | 7 (33.3) | 838 (15.0) | 1622 (11.4) | 860 (18.9) |
| 41–64 | 43 ( 45.3) | 11 (52.4) | 3243 (58.1) | 6431 (45.0) | 2124 (46.7) |
| 65-80 | 31 (32.6) | 3 (14.3) | 1503 (26.9) | 6227 (43.6) | 1560 (34.3) |
| **Sex** | | | | | |
| Men | 35 (36.8) | 9 (42.9) | 2070 (37.1) | 6436 (45.1) | 1954 (43.0) |
| Women | 60 (63.2) | 12 (57.1) | 3514 (62.9) | 7844 (54.9) | 2590 (57.0) |
| **Dynamic Lung Volumes, z-score** | | | | | |
| $FEV_1$ | −0.8 (2.0) | −1.2 (1.4) | −1. (1.7) | −0.6 (1.9) | −0.7 (1.8) |
| FVC | −0.4 (1.8) | −0.9 (1.7) | −1.3 (1.5) | −0.6 (1.9) | −0.7 (1.8) |
| $FEV_1$/FVC | −0.7 (1.8) | −0.3 (1.3) | −0.5 (1.6) | −0.6 (1.5) | −0.5 (1.4) |
| **Static Lung Volumes, z-score** | | | | | |
| TLC | −1.4 (2.0) | −2.0 (1.7) | −2.2 (1.7) | −0.9 (1.7) | −1.3 (1.8)) |
| **Interpretation** | | | | | |
| Normal | 41 (43.2) | 5 (23.8) | 1296 (23.2) | 7562 (53.0) | 2131 (46.9) |
| Non-Specific | 0 (0.0) | 2 (9.5) | 128 (2.3) | 234 (1.6) | 60 (1.3) |
| Obstructive | 18 (18.9) | 1 (4.8) | 493 (8.8) | 2253 (15.8) | 586 (12.9) |
| Restrictive | 27 (28.4) | 13 (61.9) | 3060 (54.8) | 3586 (25.1) | 1580 (34.8) |
| Mixed | 9 (9.5) | 0 (0.0) | 607 (10.9) | 645 (4.5) | 187 (4.1) |
| **Severity** | | | | | |
| Normal | 64 (67.4) | 13 (61.9) | 2986 (53.5) | 10796 (75.6) | 3456 (76.1) |
| Mild | 18 (18.9) | 6 (28.6) | 1338 (24.0) | 1847 (12.9) | 636 (14.0) |
| Moderate | 11 (11.6) | 2 (9.5) | 1105 (19.8) | 1440 (10.1) | 405 (8.9) |
| Severe | 2 (1.2) | 0 (0) | 155 (2.8) | 197 (1.4) | 47 (1.0) |

Values are median (IQR) for continuous variables and count (percentage) for categorical variables. Dynamic lung volume z-scores are calculated using GLI Global reference equations, while dynamic lung volume z-scores are calculated using GLI 2019 reference equations. Abbreviations: COPD = chronic obstructive pulmonary disease; $FEV_1$ = forced expiratory volume in 1 second; FVC = forced vital capacity; GLI = Global Lung Function Initiative; ILD = interstitial lung disease; LLN = lower limit of normal; TLC = total lung capacity.

**Table S4**. Model Performance in the Development and Validation Datasets

| **Model** | **Measure** | **Development Dataset** ($n = 42\,462$) | **Validation Dataset** ($n = 24\,524$) |
|---|---|---|---|
| FVC < LLN | Sensitivity | 0.701 (0.693 to 0.709) | 0.460 (0.450 to 0.470) |
| | Specificity | 0.836 (0.833 to 0.839) | 0.944 (0.940 to 0.947) |
| | NPV | 0.868 (0.864 to 0.872) | 0.727 (0.721 to 0.733) |
| | PPV | 0.645 (0.635 to 0.655) | 0.843 (0.833 to 0.853) |
| Logistic Regression | Sensitivity | 0.862 (0.856 to 0.867) | 0.824 (0.816 to 0.831) |
| | Specificity | 0.826 (0.823 to 0.829) | 0.835 (0.830 to 0.841) |
| | NPV | 0.934 (0.931 to 0.936) | 0.878 (0.873 to 0.884) |
| | PPV | 0.678 (0.670 to 0.685) | 0.766 (0.758 to 0.770) |
| | AUC-ROC | 0.922 (0.920 to 0.925) | 0.910 (0.906 to 0.913) |
| | AUC-PR | 0.840 (0.833 to 0.847) | 0.871 (0.871 to 0.871) |
| | ICI | 0.013 (0.011 to 0.016) | 0.190 (0.186 to 0.195) |
| | SBS | 0.520 (0.512 to 0.529) | 0.252 (0.239 to 0.265) |
| Random Forest | Sensitivity | 0.850 (0.843 to 0.858) | 0.833 (0.825 to 0.840) |
| | Specificity | 0.842 (0.838 to 0.846) | 0.829 (0.824 to 0.835) |
| | NPV | 0.930 (0.926 to 0.933) | 0.883 (0.878 to 0.889) |
| | PPV | 0.695 (0.685 to 0.706) | 0.762 (0.754 to 0.770) |
| | AUC-ROC | 0.926 (0.924 to 0.929) | 0.912 (0.909 to 0.916) |
| | AUC-PR | 0.851 (0.845 to 0.857) | 0.877 (0.877 to 0.877) |
| | ICI | 0.014 (0.011 to 0.011) | 0.172 (0.168 to 0.177) |
| | SBS | 0.532 (0.523 to 0.540) | 0.316 (0.304 to 0.329) |
| Boosted Tree | Sensitivity | 0.844 (0.838 to 0.850) | 0.830 (0.823 to 0.838) |
| | Specificity | 0.851 (0.847 to 0.855) | 0.833 (0.827 to 0.839) |
| | NPV | 0.928 (0.925 to 0.931) | 0.882 (0.877 to 0.888) |
| | PPV | 0.707 (0.697 to 0.717) | 0.766 (0.757 to 0.773) |
| | AUC-ROC | 0.928 (0.926 to 0.931) | 0.915 (0.912 to 0.919) |
| | AUC-PR | 0.856 (0.850 to 0.862) | 0.880 (0.880 to 0.880) |
| | ICI | 0.023 (0.020 to 0.026) | 0.171 (0.166 to 0.175) |
| | SBS | 0.537 (0.529 to 0.545) | 0.317 (0.305 to 0.328) |

Definition of abbreviations: AUC-PR = area under the precision recall curve; AUC-ROC = area under the receiver operating characteristic curve; FVC = forced vital capacity; ICI = integrated calibration index; LLN = lower limit of normal; NPV = negative predictive value; PPV = positive predictive value; SBS = scaled Brier score.

**Table S5**. Model Performance in the Validation Dataset by Race and Ethnicity

| Model | Measure | Asian ($n$ = 379) | Hispanic ($n$ = 61) | Non-Hispanic Black ($n$ = 11 421) | Non-Hispanic White ($n$ = 27 374) | Other ($n$ = 6 084) |
|---|---|---|---|---|---|---|
| FVC < LLN | Sensitivity | 0.472 (0.314 to 0.645) | 0.462 (0.200 to 0.750) | 0.536 (0.520 to 0.551) | 0.423 (0.520 to 0.438) | 0.392 (0.371 to 0.415) |
| | Specificity | 0.949 (0.891 to 1.000) | 0.875 (0.571 to 1.000) | 0.872 (0.857 to 0.885) | 0.951 (0.857 to 0.954) | 0.969 (0.962 to 0.975) |
| | NPV | 0.747 (0.649 to 0.849) | 0.500 (0.231 to 0.769) | 0.495 (0.478 to 0.512) | 0.796 (0.478 to 0.803) | 0.715 (0.701 to 0.730) |
| | PPV | 0.850 (0.667 to 1.000) | 0.857 (0.500 to 1.000) | 0.889 (0.876 to 0.900) | 0.783 (0.876 to 0.798) | 0.888 (0.866 to 0.910) |
| Logistic Regression | Sensitivity | 0.806 (0.667 to 0.923) | 0.923 (0.750 to 1.000) | 0.869 (0.858 to 0.880) | 0.787 (0.858 to 0.799) | 0.817 (0.799 to 0.834) |
| | Specificity | 0.881 (0.797 to 0.962) | 0.625 (0.273 to 1.000) | 0.665 (0.644 to 0.686) | 0.861 (0.644 to 0.868) | 0.860 (0.847 to 0.872) |
| | NPV | 0.881 (0.800 to 0.952) | 0.833 (0.500 to 1.000) | 0.726 (0.706 to 0.749) | 0.906 (0.706 to 0.911) | 0.881 (0.870 to 0.892) |
| | PPV | 0.806 (0.667 to 0.933) | 0.800 (0.583 to 1.000) | 0.832 (0.821 to 0.843) | 0.704 (0.821 to 0.717) | 0.788 (0.769 to 0.806) |
| | AUC-ROC | 0.894 (0.830 to 0.955) | 0.865 (0.673 to 1.000) | 0.860 (0.849 to 0.870) | 0.909 (0.849 to 0.914) | 0.916 (0.908 to 0.924) |
| | AUC-PR | 0.815 (0.669 to 0.900) | 0.839 (0.596 to 0.923) | 0.914 (0.905 to 0.923) | 0.826 (0.905 to 0.837) | 0.876 (0.861 to 0.891) |
| | ICI | 0.184 (0.118 to 0.269) | 0.319 (0.163 to 0.485) | 0.305 (0.294 to 0.316) | 0.143 (0.294 to 0.148) | 0.199 (0.188 to 0.209) |
| | SBS | 0.213 (0.013 to 0.415) | -0.007 (-1.239 to 0.507) | -0.156 (-0.206 to -0.106) | 0.301 (-0.206 to 0.316) | 0.234 (0.204 to 0.264) |
| Random Forest | Sensitivity | 0.750 (0.590 to 0.892) | 0.846 (0.615 to 1.000) | 0.881 (0.870 to 0.892) | 0.797 (0.870 to 0.809) | 0.817 (0.799 to 0.837) |
| | Specificity | 0.864 (0.770 to 0.944) | 0.750 (0.429 to 1.000) | 0.668 (0.648 to 0.688) | 0.852 (0.648 to 0.858) | 0.860 (0.847 to 0.873) |
| | NPV | 0.850 (0.754 to 0.933) | 0.750 (0.400 to 1.000) | 0.746 (0.725 to 0.766) | 0.909 (0.725 to 0.915) | 0.881 (0.868 to 0.894) |
| | PPV | 0.771 (0.622 to 0.903) | 0.846 (0.643 to 1.000) | 0.835 (0.824 to 0.847) | 0.694 (0.824 to 0.706) | 0.788 (0.770 to 0.806) |
| | AUC-ROC | 0.892 (0.817 to 0.954) | 0.875 (0.650 to 1.000) | 0.869 (0.860 to 0.879) | 0.910 (0.860 to 0.915) | 0.919 (0.911 to 0.926) |
| | AUC-PR | 0.819 (0.660 to 0.905) | 0.829 (0.567 to 0.933) | 0.922 (0.915 to 0.930) | 0.831 (0.915 to 0.841) | 0.883 (0.871 to 0.895) |
| | ICI | 0.183 (0.114 to 0.260) | 0.276 (0.132 to 0.460) | 0.279 (0.269 to 0.288) | 0.128 (0.269 to 0.133) | 0.181 (0.170 to 0.192) |
| | SBS | 0.270 (0.051 to 0.463) | -0.028 (-1.178 to 0.489) | -0.037 (-0.083 to 0.005) | 0.352 (-0.083 to 0.368) | 0.302 (0.273 to 0.334) |
| Boosted Tree | Sensitivity | 0.778 (0.630 to 0.906) | 0.846 (0.625 to 1.000) | 0.882 (0.871 to 0.892) | 0.795 (0.871 to 0.807) | 0.808 (0.789 to 0.826) |
| | Specificity | 0.864 (0.774 to 0.941) | 0.750 (0.400 to 1.000) | 0.663 (0.643 to 0.684) | 0.856 (0.643 to 0.863) | 0.867 (0.854 to 0.879) |
| | NPV | 0.864 (0.774 to 0.947) | 0.750 (0.400 to 1.000) | 0.745 (0.726 to 0.765) | 0.909 (0.726 to 0.914) | 0.876 (0.864 to 0.889) |
| | PPV | 0.778 (0.639 to 0.903) | 0.846 (0.611 to 1.000) | 0.833 (0.821 to 0.844) | 0.700 (0.821 to 0.713) | 0.794 (0.775 to 0.813) |
| | AUC-ROC | 0.899 (0.828 to 0.957) | 0.865 (0.645 to 1.000) | 0.870 (0.860 to 0.879) | 0.914 (0.860 to 0.918) | 0.923 (0.915 to 0.931) |
| | AUC-PR | 0.820 (0.665 to 0.909) | 0.836 (0.585 to 0.924) | 0.922 (0.914 to 0.929) | 0.834 (0.914 to 0.843) | 0.888 (0.875 to 0.901) |
| | ICI | 0.170 (0.119 to 0.259) | 0.301 (0.177 to 0.486) | 0.279 (0.268 to 0.289) | 0.125 (0.268 to 0.131) | 0.182 (0.171 to 0.194) |
| | SBS | 0.270 (0.064 to 0.460) | 0.001 (-1.315 to 0.473) | -0.038 (-0.082 to 0.005) | 0.353 (-0.082 to 0.368) | 0.301 (0.271 to 0.328) |

Definition of abbreviations: AUC-PR = area under the precision recall curve; AUC-ROC = area under the receiver operating characteristic curve; FVC = forced vital capacity; ICI = integrated calibration index; LLN = lower limit of normal; NPV = negative predictive value; PPV = positive predictive value; SBS = scaled Brier score.

**Table S6**. Model Performance in the Validation Dataset by Age Group

| Model | Measure | Age < 65 ($n = 15\,200$) | Age ≥ 65 ($n = 9\,324$) |
|---|---|---|---|
| FVC < LLN | Sensitivity | 0.468 (0.440 to 0.496) | 0.459 (0.448 to 0.470) |
| | Specificity | 0.972 (0.965 to 0.979) | 0.939 (0.934 to 0.943) |
| | NPV | 0.775 (0.759 to 0.791) | 0.719 (0.713 to 0.726) |
| | PPV | 0.899 (0.875 to 0.923) | 0.835 (0.824 to 0.847) |
| Logistic Regression | Sensitivity | 0.840 (0.831 to 0.849) | 0.794 (0.781 to 0.806) |
| | Specificity | 0.842 (0.834 to 0.849) | 0.825 (0.815 to 0.835) |
| | NPV | 0.882 (0.875 to 0.889) | 0.873 (0.865 to 0.882) |
| | PPV | 0.789 (0.779 to 0.799) | 0.725 (0.711 to 0.738) |
| | AUC-ROC | 0.919 (0.914 to 0.923) | 0.893 (0.887 to 0.900) |
| | AUC-PR | 0.888 (0.881 to 0.895) | 0.838 (0.826 to 0.850) |
| | ICI | 0.196 (0.190 to 0.202) | 0.181 (0.174 to 0.189) |
| | SBS | 0.259 (0.242 to 0.275) | 0.236 (0.215 to 0.257) |
| Random Forest | Sensitivity | 0.843 (0.835 to 0.852) | 0.812 (0.799 to 0.825) |
| | Specificity | 0.839 (0.832 to 0.847) | 0.814 (0.804 to 0.825) |
| | NPV | 0.884 (0.877 to 0.890) | 0.882 (0.874 to 0.891) |
| | PPV | 0.787 (0.778 to 0.797) | 0.718 (0.702 to 0.732) |
| | AUC-ROC | 0.922 (0.918 to 0.926) | 0.896 (0.889 to 0.902) |
| | AUC-PR | 0.895 (0.889 to 0.902) | 0.842 (0.829 to 0.852) |
| | ICI | 0.178 (0.172 to 0.184) | 0.164 (0.157 to 0.172) |
| | SBS | 0.333 (0.318 to 0.349) | 0.285 (0.265 to 0.305) |
| Boosted Tree | Sensitivity | 0.841 (0.831 to 0.850) | 0.811 (0.797 to 0.824) |
| | Specificity | 0.850 (0.842 to 0.858) | 0.808 (0.798 to 0.817) |
| | NPV | 0.883 (0.877 to 0.890) | 0.880 (0.872 to 0.889) |
| | PPV | 0.798 (0.787 to 0.808) | 0.710 (0.696 to 0.723) |
| | AUC-ROC | 0.925 (0.921 to 0.929) | 0.898 (0.892 to 0.904) |
| | AUC-PR | 0.898 (0.891 to 0.904) | 0.843 (0.832 to 0.853) |
| | ICI | 0.177 (0.172 to 0.183) | 0.160 (0.153 to 0.167) |
| | SBS | 0.330 (0.315 to 0.346) | 0.289 (0.270 to 0.308) |

Definition of abbreviations: AUC-PR = area under the precision recall curve; AUC-ROC = area under the receiver operating characteristic curve; FVC = forced vital capacity; ICI = integrated calibration index; LLN = lower limit of normal; NPV = negative predictive value; PPV = positive predictive value; SBS = scaled Brier score.

**Table S7**. Model Performance in the Validation Dataset by Sex

| **Model** | **Measure** | **Male** ($n = 10\,504$) | **Female** ($n = 14\,020$) |
|---|---|---|---|
| FVC < LLN | Sensitivity | 0.498 (0.483 to 0.513) | 0.430 (0.419 to 0.444) |
| | Specificity | 0.933 (0.927 to 0.939) | 0.951 (0.947 to 0.956) |
| | NPV | 0.732 (0.722 to 0.742) | 0.724 (0.716 to 0.732) |
| | PPV | 0.836 (0.822 to 0.850) | 0.849 (0.836 to 0.863) |
| Logistic Regression | Sensitivity | 0.852 (0.841 to 0.862) | 0.802 (0.791 to 0.813) |
| | Specificity | 0.821 (0.810 to 0.830) | 0.846 (0.838 to 0.853) |
| | NPV | 0.890 (0.882 to 0.898) | 0.870 (0.863 to 0.877) |
| | PPV | 0.764 (0.751 to 0.776) | 0.768 (0.757 to 0.780) |
| | AUC-ROC | 0.919 (0.913 to 0.924) | 0.904 (0.899 to 0.909) |
| | AUC-PR | 0.887 (0.887 to 0.887) | 0.860 (0.860 to 0.860) |
| | ICI | 0.165 (0.158 to 0.172) | 0.210 (0.204 to 0.216) |
| | SBS | 0.337 (0.315 to 0.356) | 0.187 (0.170 to 0.204) |
| Random Forest | Sensitivity | 0.864 (0.853 to 0.875) | 0.808 (0.797 to 0.818) |
| | Specificity | 0.802 (0.791 to 0.812) | 0.850 (0.842 to 0.857) |
| | NPV | 0.896 (0.888 to 0.904) | 0.874 (0.867 to 0.881) |
| | PPV | 0.748 (0.736 to 0.760) | 0.774 (0.763 to 0.785) |
| | AUC-ROC | 0.919 (0.914 to 0.924) | 0.909 (0.904 to 0.914) |
| | AUC-PR | 0.891 (0.891 to 0.891) | 0.867 (0.867 to 0.867) |
| | ICI | 0.152 (0.145 to 0.159) | 0.187 (0.181 to 0.194) |
| | SBS | 0.380 (0.361 to 0.398) | 0.268 (0.251 to 0.284) |
| Boosted Tree | Sensitivity | 0.857 (0.848 to 0.868) | 0.809 (0.799 to 0.819) |
| | Specificity | 0.812 (0.802 to 0.823) | 0.848 (0.841 to 0.856) |
| | NPV | 0.893 (0.885 to 0.901) | 0.874 (0.867 to 0.882) |
| | PPV | 0.757 (0.745 to 0.770) | 0.773 (0.762 to 0.783) |
| | AUC-ROC | 0.921 (0.915 to 0.926) | 0.912 (0.908 to 0.917) |
| | AUC-PR | 0.893 (0.893 to 0.893) | 0.870 (0.870 to 0.870) |
| | ICI | 0.151 (0.144 to 0.158) | 0.186 (0.180 to 0.192) |
| | SBS | 0.381 (0.364 to 0.398) | 0.267 (0.251 to 0.283) |

Definition of abbreviations: AUC-PR = area under the precision recall curve; AUC-ROC = area under the receiver operating characteristic curve; FVC = forced vital capacity; ICI = integrated calibration index; LLN = lower limit of normal; NPV = negative predictive value; PPV = positive predictive value; SBS = scaled Brier score.

**Figure S1**. Flow Diagram for Inclusion in the Development Dataset

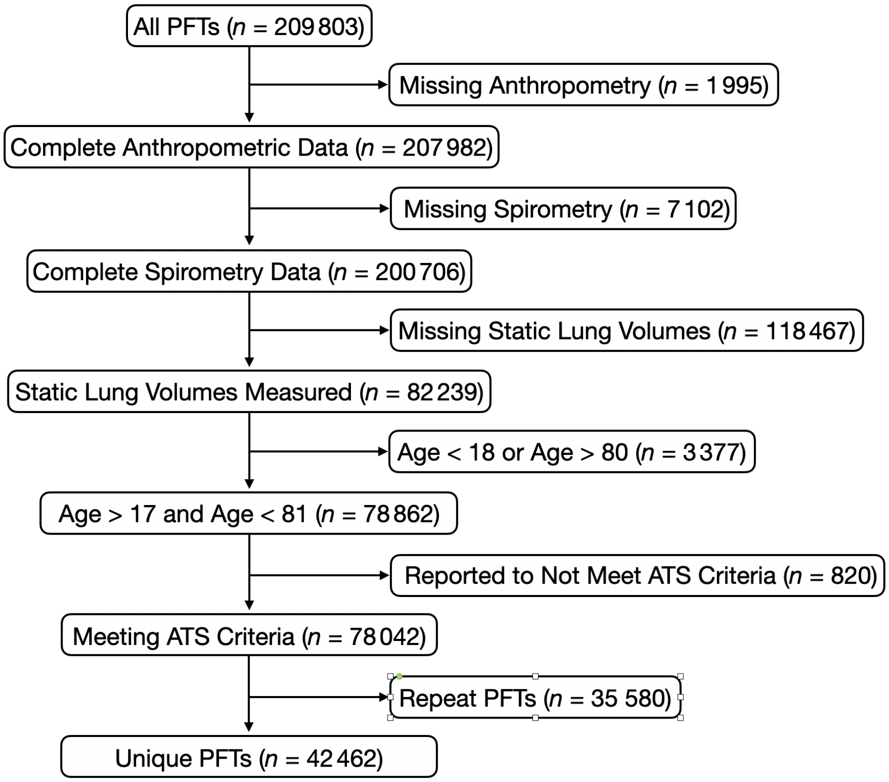

**Figure S2**. Forced Vital Capacity Z-Score and Negative Predictive Value

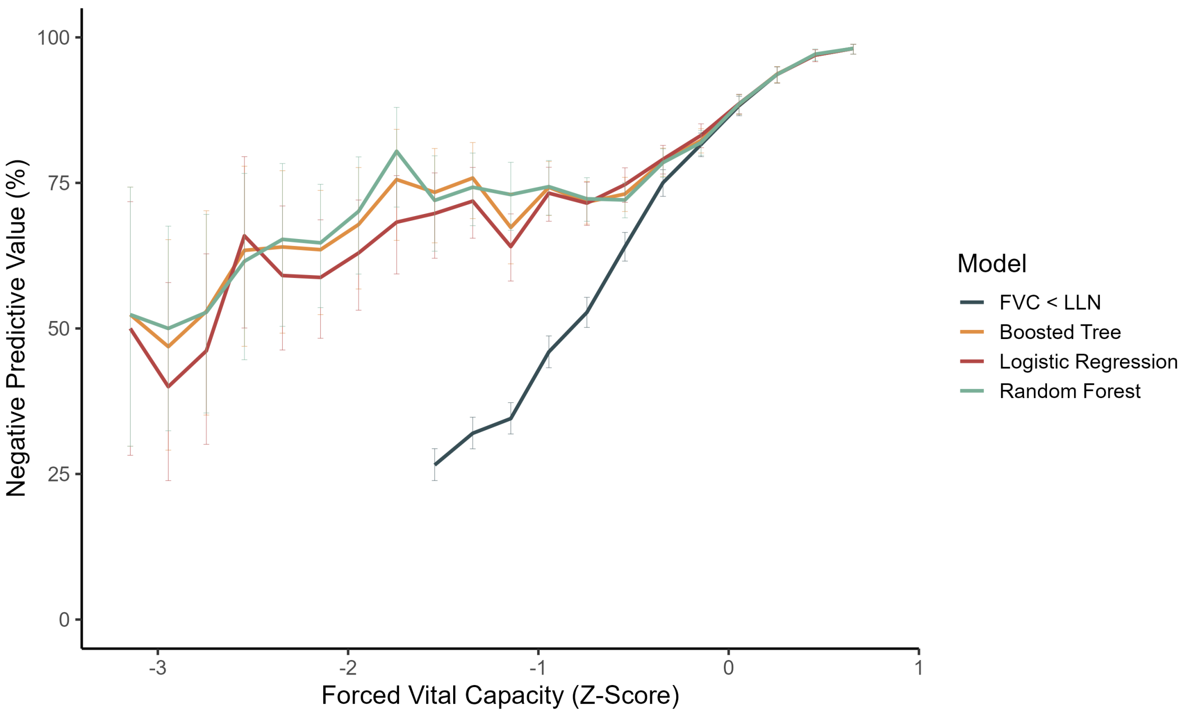

**Figure S3**. Forced Vital Capacity Z-Score and the Exclusion of Restriction

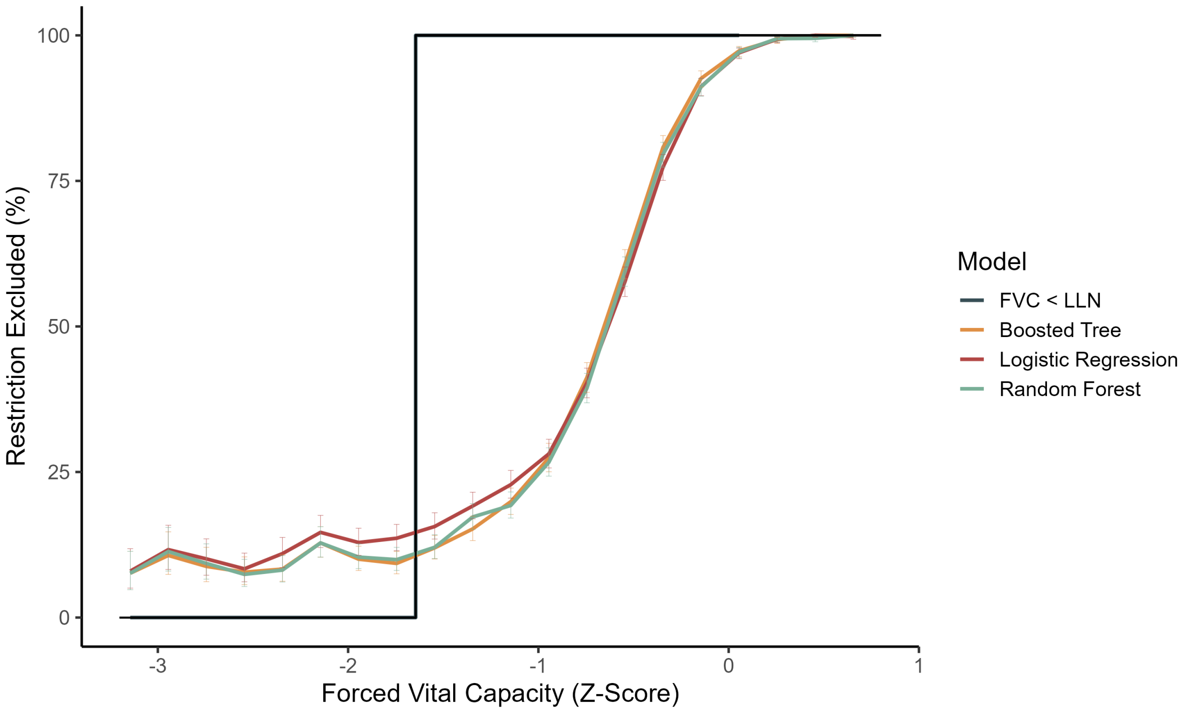

**Figure S4**. Boosted Tree Model Shapley Values

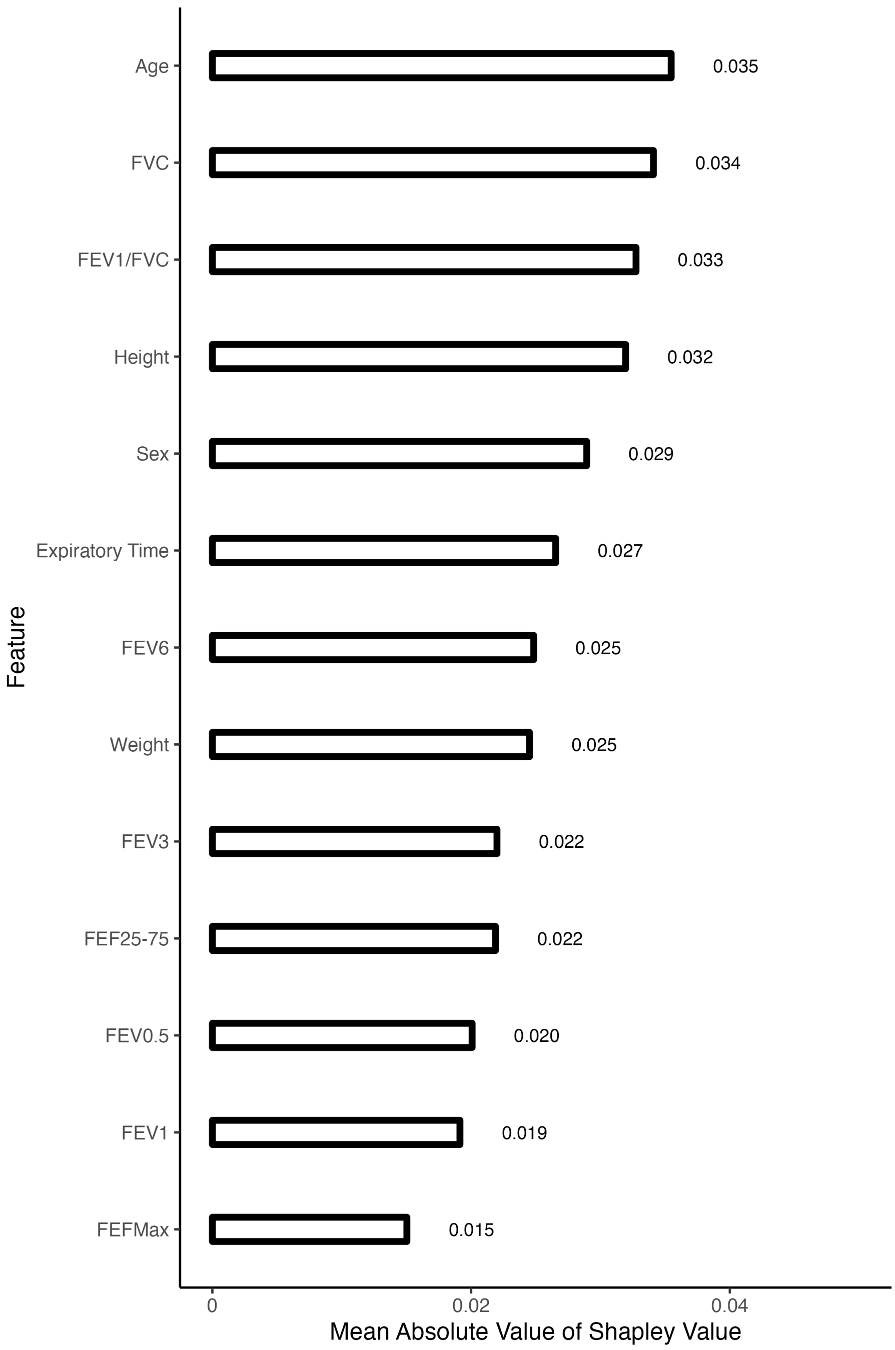

**Figure S5**. Logistic Regression Model Shapley Values

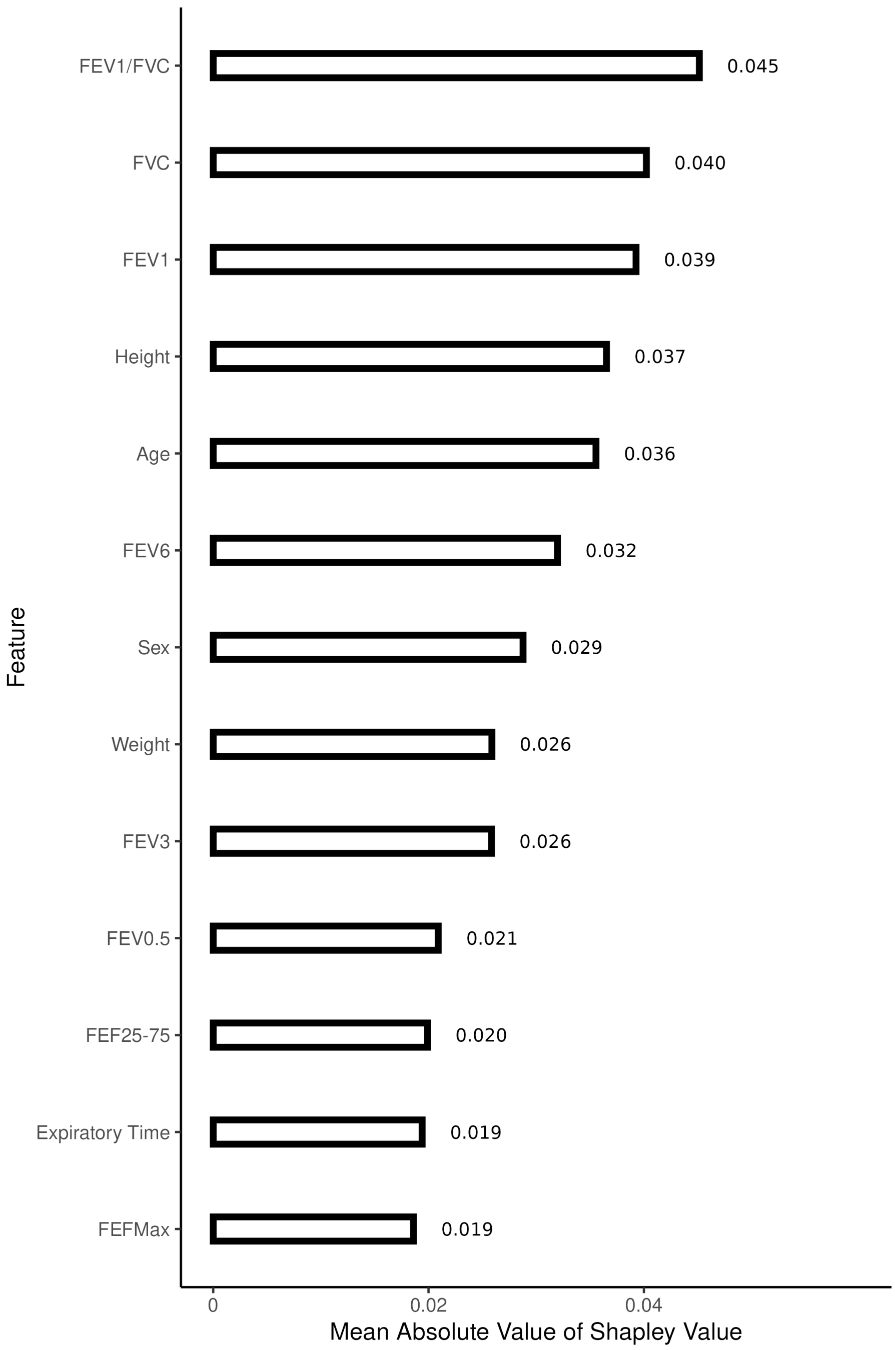

**Figure S6**. Random Forest Model Shapley Values

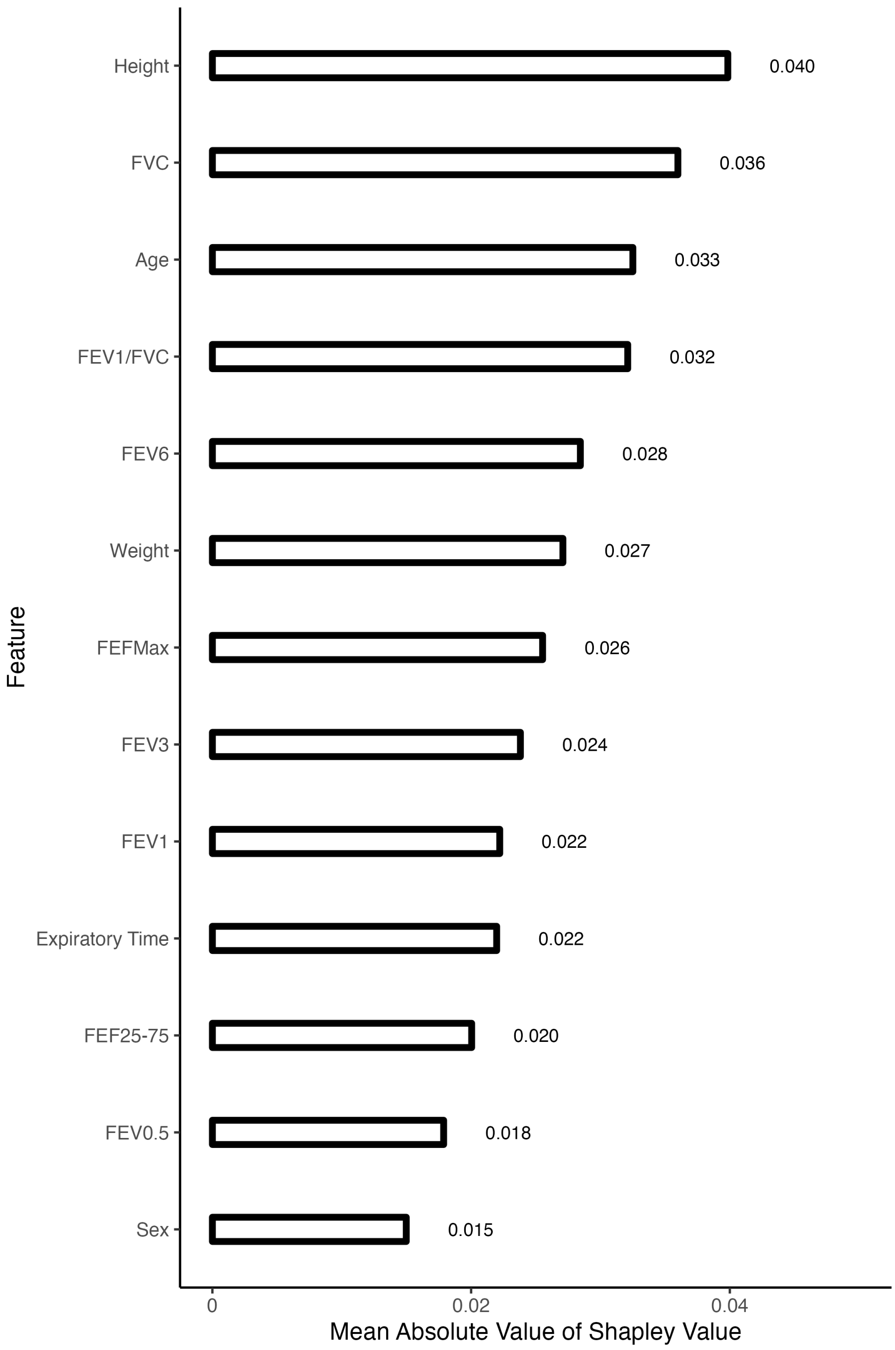
